# Blind validation of MSIntuit, an AI-based pre-screening tool for MSI detection from histology slides of colorectal cancer

**DOI:** 10.1101/2022.11.17.22282460

**Authors:** Charlie Saillard, Rémy Dubois, Oussama Tchita, Nicolas Loiseau, Thierry Garcia, Aurélie Adriansen, Séverine Carpentier, Joelle Reyre, Diana Enea, Aurélie Kamoun, Stéphane Rossat, Meriem Sefta, Michael Auffret, Lionel Guillou, Arnaud Fouillet, Jakob Nikolas Kather, Magali Svrcek

## Abstract

**Objective:** Mismatch Repair Deficiency (dMMR) / Microsatellite Instability (MSI) is a key biomarker in colorectal cancer (CRC). Universal screening of CRC patients for dMMR/MSI status is now recommended, but contributes to increased workload for pathologists and delayed therapeutic decisions. Deep learning has the potential to ease dMMR/MSI testing in clinical practice, yet no comprehensive validation of a clinically approved tool has been conducted.

**Design:** We developed an MSI pre-screening tool, MSIntuit, that uses deep learning to identify MSI status from H&E slides. For training, we used 859 slides from the TCGA database. A blind validation was subsequently performed on an independent dataset of 600 consecutive CRC patients. Each slide was digitised using Phillips-UFS and Ventana-DP200 scanners. Thirty dMMR/MSI slides were used for calibration on each scanner. Prediction was then performed on the remaining 570 patients following an automated quality check step. The inter and intra-scanner reliability was studied to assess MSIntuit’s robustness.

**Results:** MSIntuit reached a sensitivity and specificity of 97% (95% CI: 93-100%) / 46% (42-50%) on DP200 and of 95% (90-98%) / 47% (43-51%) on UFS scanner. MSIntuit reached excellent agreement on the two scanners (Cohen’s κ: 0.82) and was repeatable across multiple rescanning of the same slide (Fleiss’ κ: 0.82).

**Conclusion:** We performed a successful blind validation of the first clinically approved AI-based tool for MSI detection from H&E slides. MSIntuit reaches sensitivity comparable to gold standard methods (92-95%) while ruling out almost half of the non-MSI population, paving the way for its use in clinical practice.

## INTRODUCTION

Microsatellite Instability (MSI) is a tumour genotype characterised by mismatch errors of repetitive DNA sequences, called microsatellites, distributed along the genome. It is caused by a deficiency in the DNA mismatch repair (MMR) system, the process whereby errors, such as insertions and deletions, that occur during DNA replication are recognized and fixed. MSI occurs due to MMR malfunction and is therefore a marker of mismatch repair deficiency (dMMR). dMMR tumours result from defects in the major MMR genes, namely MLH1, MSH2, MSH6 and PMS2. MSI is found in approximately 15% of the colorectal cancer (CRC) population and plays a crucial role in clinical management of CRC, with major diagnostic, prognostic and therapeutic implications. First, MSI is the hallmark of Lynch Syndrome (LS), the most frequent form of hereditary predisposition to develop CRC. Second, dMMR/MSI is associated with better prognosis in early-stage CRC and a lack of benefit from adjuvant treatment with 5-fluorouracil in stage II disease. Third, dMMR/MSI tumours are sensitive to immune checkpoint inhibitor treatments. In 2017, this genomic instability phenotype became the first pan-cancer biomarker approved by the FDA, allowing the use of pembrolizumab for patients with dMMR/MSI unresectable or metastatic solid tumours [1]. Given all the implications of MSI in patient care, many medical organisations (such as NICE and NCCN [2,3]) recommend universal screening for dMMR/MSI status of newly diagnosed CRC.

dMMR/MSI can be diagnosed with immunohistochemistry (MMR-IHC) to detect loss of MMR proteins and/or by molecular tests such as polymerase chain reaction (MSI-PCR) to show microsatellite alterations. Next Generation Sequencing (NGS) represents an alternative molecular test to diagnose MSI. MMR-IHC and MSI-PCR testing are relatively inexpensive and well-established methods to detect molecular alterations in developed countries, while NGS is too expensive to be used in clinical routine. MMR-IHC testing requires excellent tissue fixation, slide preparation time, an experienced pathologist and consumes tissue material which can be in very limited supply for small tumours where multiple tests need to be performed. It also lacks standardisation across labs and can be subject to inter-rater (pathologist) variability. MSI-PCR testing requires specific infrastructure that may not be available in every centre and has a longer turnaround time which can delay therapeutic decisions. As the number of biomarkers has steadily increased over the last two decades [4], MMR-IHC and MSI-PCR testing contribute to an ever-increasing workload for pathologists and technicians. Given the global shortage of pathologists worldwide [5], leveraging AI could ease the testing burden of MSI by reducing the workload of pathologists, shortening delay in therapeutic decisions, decreasing costs and preserving material. In a 2019 study, we showed that deep learning could accurately detect MSI from H&E slides in CRC [6]. Since then, several studies have presented deep learning-based MSI classifiers from H&E slides in CRC, confirming its potential to complement standard MSI screening methods [7–9].

Despite recent advances, several issues remain that prevent AI-based tools for MSI prediction from being used in clinical practice, as pointed out by Kleppe [10]. Most existing studies focus on the area under the ROC curve (AUROC) as their main performance metric. Although useful to compare performance of several machine learning models, this metric can hide a severe lack of generalisation and is not relevant to clinical practice. Instead, we propose to focus on sensitivity and specificity to evaluate diagnostic accuracy of MSIntuit™ CRC (MSIntuit), an AI enabled pre-screening solution that enables an early rule-out of non-MSI patients using H&E slides from primary colorectal tumour by outputting two categories (Undetermined/MSS-AI) (figure 1A). Importantly, an MSI pre-screening tool used as a rule-out test must have a very high sensitivity. We therefore propose a method that guarantees the sensitivity is maintained at new sites and on new cohorts.

**Figure 1.**
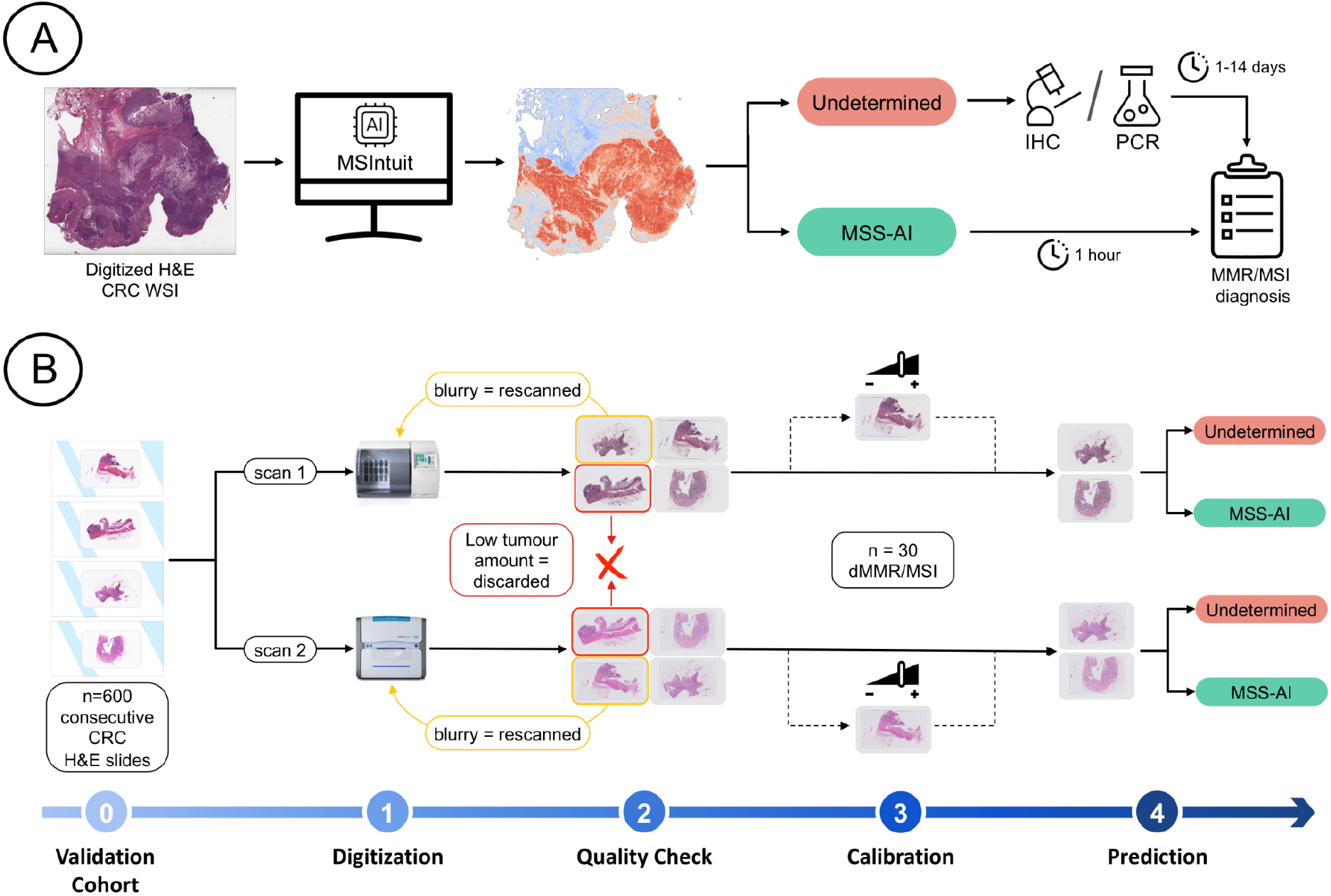
Clinical workflow and blind validation methodology. **A)** Clinical workflow of MSI screening with MSIntuit. Using a routine H&E slide of CRC, MSIntuit outputs if the patient is likely to be MSI (Undetermined) and should receive a confirmatory test (MMR-IHC and/or MSI-PCR), or not (MSS-AI). By ruling out a significant fraction of non-MSI patients, the workload of pathologists is reduced and the MSI screening is fastened. **B)** H&E slides of 600 consecutive resected CRC specimens were collected and digitised on two scanners, Phillips UFS and Ventana DP200, resulting in two sets of slides: MPATH-UFS and MPATH-DP200 (step 1). For each cohort, the same pipeline was then applied: an automated quality check discarded slides that did not match criteria (large blurry regions, too few tumour). Slides with large blurry regions were rescanned (step 2). Next, 30 dMMR/MSI WSIs were selected randomly and used to define an appropriate threshold (step 3). Finally, MSIntuit prediction was performed on the remaining slides using the threshold defined in the aforementioned step to classify patients into two categories: MSS-AI and Undetermined (step 4).

To our knowledge, no studies evaluating performance of tools based on deep learning models to predict MSI from histology slides have solved the issue of model generalisability in such a way as to enable its use in clinical routine. In this study, we performed a blind clinical validation of MSIntuit on a large external cohort of 600 consecutive CRC cases. We find that used as a pre-screening tool, MSIntuit can rule out almost half of the non-MSI population to ease MSI screening. Our tool includes an automatic slide quality check and addresses the issue of defining an operating threshold with a calibration step, making it directly applicable to clinical practice. We further studied MSIntuit’s robustness to potential sources of variability at new sites by 1) digitising the 600 same slides with two different scanners (inter-scanner reliability), and 2) digitising 30 slides 8 times on the same scanner (intra-scanner reliability). For a subset of 200 tumours, we also studied the impact of slide selection on MSIntuit performance by collecting 1 to 4 additional slides from different blocks of the same tumour (inter-block reliability). Taken together, these studies demonstrate the analytical robustness of MSIntuit, and pave the way to its adoption in clinical routine.

## MATERIALS & METHODS

### Cohort description

Three cohorts were used in our study: a discovery cohort to train our model, an independent development cohort to gain insights about the model performances on an external dataset, and an independent validation cohort, blinded to patients’ MSI statuses, to assess the performance of MSIntuit in a one-shot fashion. Inclusion criteria for all cohorts were as follows: unequivocal diagnosis of CRC, available histological slides of resected specimens from the primary tumour, available MSI status. The discovery cohort, denoted TCGA here, is a multicentric cohort of 859 whole slide images (WSI) from 434 patients from the TCGA-COAD database diagnosed in 24 US centres [11]. 427 Formalin-Fixed Paraffin-Embedded (FFPE) and 432 snap frozen H&E-stained WSIs from these patients were used to develop our model. MSI-PCR was used as a source of ground truth for MSI labels. The PAIP cohort was used as a development set and comprised colorectal tumour samples of n=47 patients, collected from three centres in South Korea [12]. The MSI status of these patients was determined using MSI-PCR assays. The validation cohort used for the blind validation consisted of 600 anonymized FFPE H&E WSIs of 600 consecutive patients diagnosed at Medipath pathology laboratories (France) in 2017 and 2018. For each patient, one H&E slide was chosen following our guidelines (see *Slide selection* paragraph below). All slides were digitised using two scanners, Philips UFS (Philips, Amsterdam, The Netherlands) and Ventana DP200 (Roche Diagnostics GmbH, Mannheim, Germany), leading to two sets of 600 WSIs referred to as MPATH-UFS and MPATH-DP200. dMMR status was assessed using MMR-IHC for the four MMR proteins, and confirmed by MSI-PCR for n=33 indeterminate cases (doubt in MMR-IHC interpretation or suspicion of Lynch Syndrome). Clinicopathologic features of these three cohorts can be found in online supplemental table 1.

### Performance metrics

This study is reported in accordance with the Standards for Reporting of Diagnostic Accuracy Studies guidelines [13]. The clinical value of the models was evaluated using sensitivity, specificity and negative predictive value (NPV) metrics. Raw performance of the models was also evaluated using the area under the receiver operating characteristic curve (AUROC). Confidence intervals were generated using bootstrapping with 1000 repetitions.

### Inter and intra-scanner reliability analysis

The intraclass Correlation Coefficients (ICC) was used to measure the agreement of the continuous predictions of the same slides digitised with UFS and DP200 scanners. Specifically, we used a single-measurement (i.e. same patient), absolute agreement, two-way mixed effects (fixed raters i.e. scanners across all targets i.e. patients) model which corresponds to the ICC(A, 2) form [14]. The ICC value indicates how much of the score variance can be explained by random effects (subjects) and not fixed effects (scanners). Cohen’s kappa statistic was used to assess the agreement between MSIntuit’s predicted classes across the two scanners and the patients’ MSI status. Fleiss’ Kappa statistic was used to study the agreement of MSIntuit’s predicted classes for the same slides digitised eight times and patient’s MSI status.

### Automated quality check

Automated quality check (QC) consisted of two steps: detection of large artefact regions and detection of tumour regions (figure 1B). For the first step, artefact regions such as blurry areas were discarded via a UNet trained to detect tissue on the slides. Details about this neural network can be found in online supplemental methods. The tissue mask generated by the UNet was then briefly examined by a technician to identify artefacts, potentially leading to a new digitization of the slide. For the second step, a tumour detection model was applied to determine which tiles were tumoral and which tiles were not. This model was a multilayer perceptron (MLP) with one hidden layer of 256 neurons with ReLU activation, that was trained with MoCo features (online supplemental methods) of 642,122 tiles from 50 slides of TCGA-COAD, based on tumour annotations made by an expert pathologist. A minimum number of 500 tumour tiles, which corresponds to approximately 6 mm^2^, was set as the cut-off to pass QC, based on empirical evidence obtained from the development cohorts (online supplemental figure 1)

### Model description

Model used to predict MSI status from slide features was a variant of Chowder [15]. Details about feature extraction are provided in the online supplemental methods. First layer of Chowder was an MLP with 128 hidden neurons and sigmoid activation that was applied to each tile’s features to output one score. The 10 top and bottom scores were then concatenated and fed into a MLP with 128 and 64 hidden neurons and sigmoid activations. The model was trained with binary cross entropy as loss, with weights balanced with respect to the prevalence of dMMR/MSI in the discovery set.

### Tool’s consistency across slides from different blocks of the same tumour

For a subset of 200 patients out of the 600 patients of MPATH-DP200 dataset, 1 to 4 other tumour slides coming from different blocks of the same patient were digitised, resulting in a total of 398 additional slides. We characterise the tumour morphology of these slides (figures 4A, 4C) using a ResNet18 model trained on NCT-CRC-HE-100K dataset [16] from the TIAToolbox library [16,17]. This classifier takes as input a tile of 112 × 112 μm and outputs a probability for each of the following classes: adipose (ADI), debris (DEB), lymphocytes (LYM), mucus (MUC), smooth muscle (MUS), normal colon mucosa (NORM), cancer-associated stroma (STR), colorectal adenocarcinoma epithelium (TUM). We then assessed the variations in MSIntuit predictions according to the slide chosen to be processed by the tool. Using the tissue type classifier described above, we also determined how each tissue type category impacted MSIntuit prediction, and which kind of slide was preferable to be selected for MSIntuit processing. McNemar’s test was used to assess the significance of performance difference by selecting in each tumour, the slide with highest and lowest amount of each tissue category.

### Patient and public involvement

Patients were not involved in this study.

## RESULTS

### Quality check and calibration as preliminary steps for a clinical-ready AI-based tool

An automated quality check (QC) was first performed on MPATH-DP200 and MPATH-UFS cohorts to set aside slides that did not meet the tool requirements. This step allowed us to automatically detect slides that were not properly scanned and contained large blurry regions, which could impact the final prediction score. Interestingly, these blurry slides were not noticed by the pathologists because it was only visible at a high magnification level (online supplemental figure 2). The QC was able to identify these slides quickly, without the need for manual examination. As a result, 3% of MPATH-DP200 slides and 2% of MPATH-UFS slides were rescanned. Second step of QC allowed to detect slides with too little tumour tissue (<5mm^2^): 5% and 2% of the slides were discarded on MPATH-DP200 and MPATH-UFS cohorts, respectively.

As a result of this preprocessing, we obtained n=541 (dMMR/MSI: 87) and n=558 (dMMR/MSI: 90) slides for MPATH-DP200 and MPATH-UFS cohorts respectively.

Additionally, to address the issue of variations in data acquisition protocols such as stainers and scanners that may impact deep learning model prediction distributions, we used a calibration strategy to ensure a sensitivity between 93% and 97% was obtained for the blind validation (figure 1B). For both MPATH-DP200 and MPATH-UFS, 30 slides from the same dMMR/MSI patients were used to define the operating threshold leading to 1/30 misclassification (meaning, 1 slide was classified as MSS-AI, and 29 were classified as Undetermined). The number of slides used in this calibration step was chosen after a sensitivity analysis on several internal datasets showed that 30 slides were sufficient to ensure a high likelihood that the sensitivity of MSIntuit on the remaining samples was at least 93%. This process led us to choose a threshold of 0.20045 on the MPATH-UFS dataset and 0.20202 on the MPATH-DP200 dataset. The similarity of the two thresholds suggests that the variations between UFS and DP200 scanners did not meaningfully impact MSIntuit predictions, despite the model having been trained on data collected using another scanner (Aperio).

### MSIntuit performance was boosted using self-supervised learning, allowing it to rule out almost half of the MSS population with high sensitivity

During model development, we found that using a feature extractor pre-trained with self-supervised learning on millions of histology tiles yielded a performance improvement. We compared this approach against a method widely used in medical imaging which consists of using a feature extractor trained on the ImageNet dataset, a dataset which does not contain any histology images. Our method largely outperformed the ImageNet one with AUROCs of 0.97, 0.88, 0.86 versus 0.92 and 0.78, 0.77 for ImageNet on PAIP, MPATH-DP200 and MPATH-UFS cohorts, respectively (online supplemental figure 3A, supplemental table 2). Including frozen slides in the training set and applying our model to the whole-slide (not just the tumour content) also yielded small performance improvements (online supplemental tables 3, 4).

Following QC and calibration, predictions of MSI status were generated from the histology slides and resulted in a sensitivity of 97% (95% CI: 93-100%), a specificity of 46% (42-50%) and an NPV of 99% (97-100%) on the MPATH-DP200 cohort, and a sensitivity of 95% (90-98%), a specificity of 47% (43-51%) and an NPV of 98% (96-99%) on the MPATH-UFS cohort (figures 2A, 2B). On both cohorts, MSIntuit was therefore able to correctly identify the majority of dMMR/MSI patients while ruling out almost half of the pMMR/MSS population and enriching the remaining population to screen in dMMR/MSI patients by +63%. This shows the robustness of our calibration approach and that our model generalises well on an independent cohort and across two different scanners not used during training.

**Figure 2.**
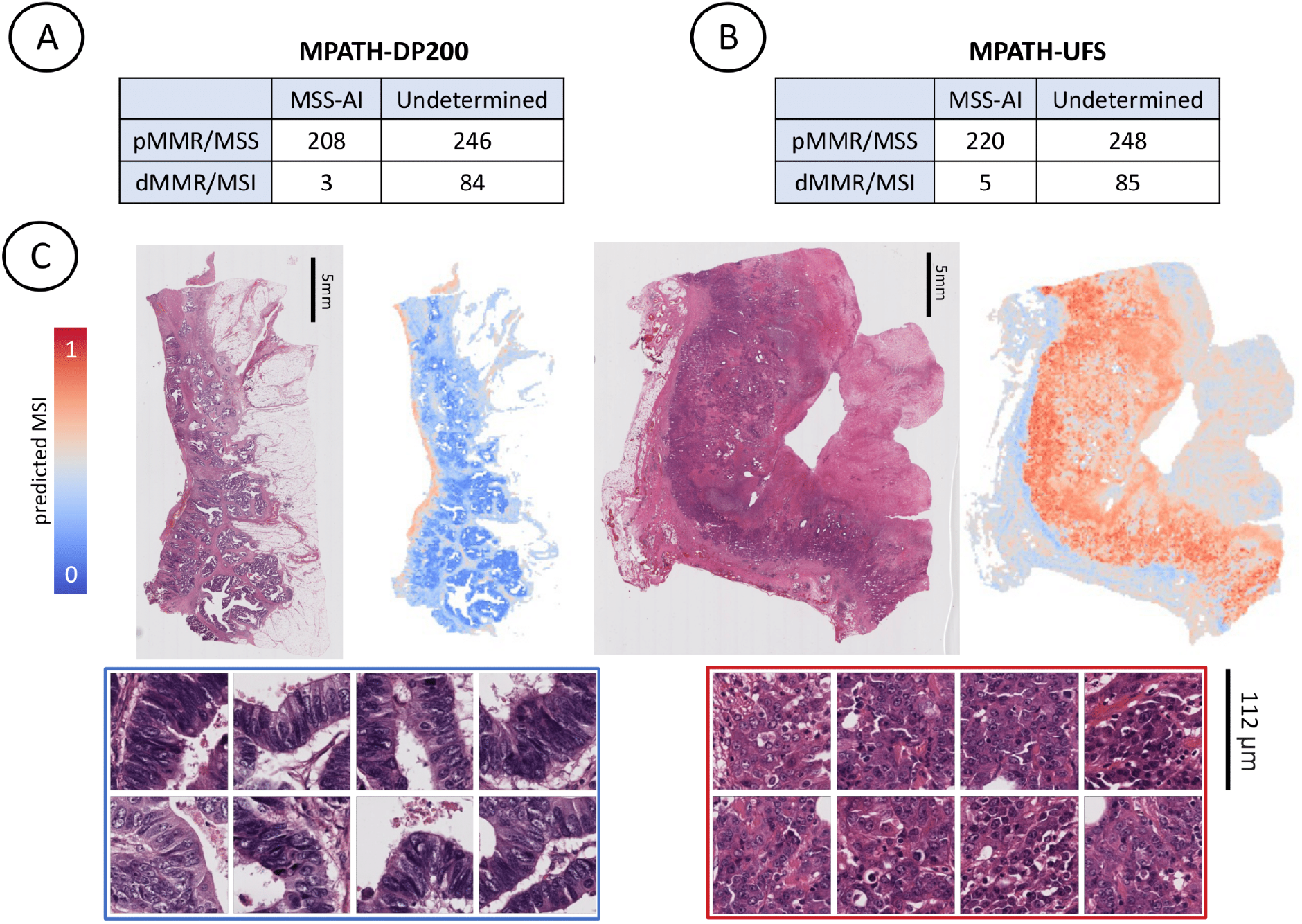
Performance and Model Interpretability.Confusion matrices of MSIntuit performance on. **A)** MPATH-DP200 cohort, **B)** MPATH-UFS cohort, **C)** Top: MSIntuit prediction heatmaps showing MSI score for each 112 × 112 μm tile on representative non-MSI (left) and MSI (right) cases. Bottom: Corresponding most predictive regions of non-MSI (left) and MSI regions (right). Regions predictive of MSI displayed poor differentiation, tumour infiltrating lymphocytes while regions predictive of non-MSI were well differentiated tumour glands.

### MSIntuit reached excellent agreement on two scanners, and is repeatable across multiple rescanning of the same slide

Several studies have shown that different scanners induce variations on the digital images generated, which can hamper the development of computational pathology (CP) tools [18,19]. Given that various scanner models are used across medical centres, it is crucial that CP tools can handle these data acquisition variabilities. Results presented in the previous section show that MSIntuit generalises well to scanners not used during model training. To further study this potential issue, we assessed the impact of digitization variations on MSIntuit by comparing the results obtained on MPATH-DP200 and MPATH-UFS cohorts, which were composed of the exact same slides digitised with these two different scanners. We first compared the results obtained on the exact same set of slides across the two scanners, and found out that model performance was extremely close: AUROCs of 0.87 (95% CI: 0.84-0.90) and 0.87 (0.84-0.91) were obtained on DP200 and UFS scanners respectively (figure 3A). Additionally, correlation of predictions across the two scanners was very strong with a Pearson’s R of 0.98 (p<0.001, figure 3B). Interestingly, the correlation was substantially lower using an ImageNet pre-trained feature extractor (R=0.82, online supplementary figure 3B). Since Pearson’s correlation is a good measure of linear correlation but not of absolute agreement, we also computed the intra-class correlation coefficient (ICC): inter-scanner reliability was excellent with an ICC of 0.99 (95% CI: 0.99-0.99). We also measured the agreement of the categories outputted by MSIntuit on the two scanners: an almost perfect agreement was observed with a Cohen’s Kappa of 0.82. As MSIntuit also outputs one score per tile (representing the likelihood of the tile belonging to a dMMR/MSI slide), we also assessed the model’s robustness to the scanner at this finer level (figure 3D). 272,527 tiles of 20 slides sampled randomly (dMMR/MSI: 10, non-dMMR/MSI: 10) were used and a score was generated for each of them on the two scanners. A very strong correlation was observed with a Pearson’s R of 0.92 (p<0.001, figure 3E). Finally, we assessed the intra-scanner reliability of our tool by looking at the process of digitization: 30 slides were digitised 8 times on the UFS scanner. Agreement of the tool across the different digitizations was almost perfect with a Fleiss’ Kappa of 0.82 (figure 3C).

**Figure 3.**
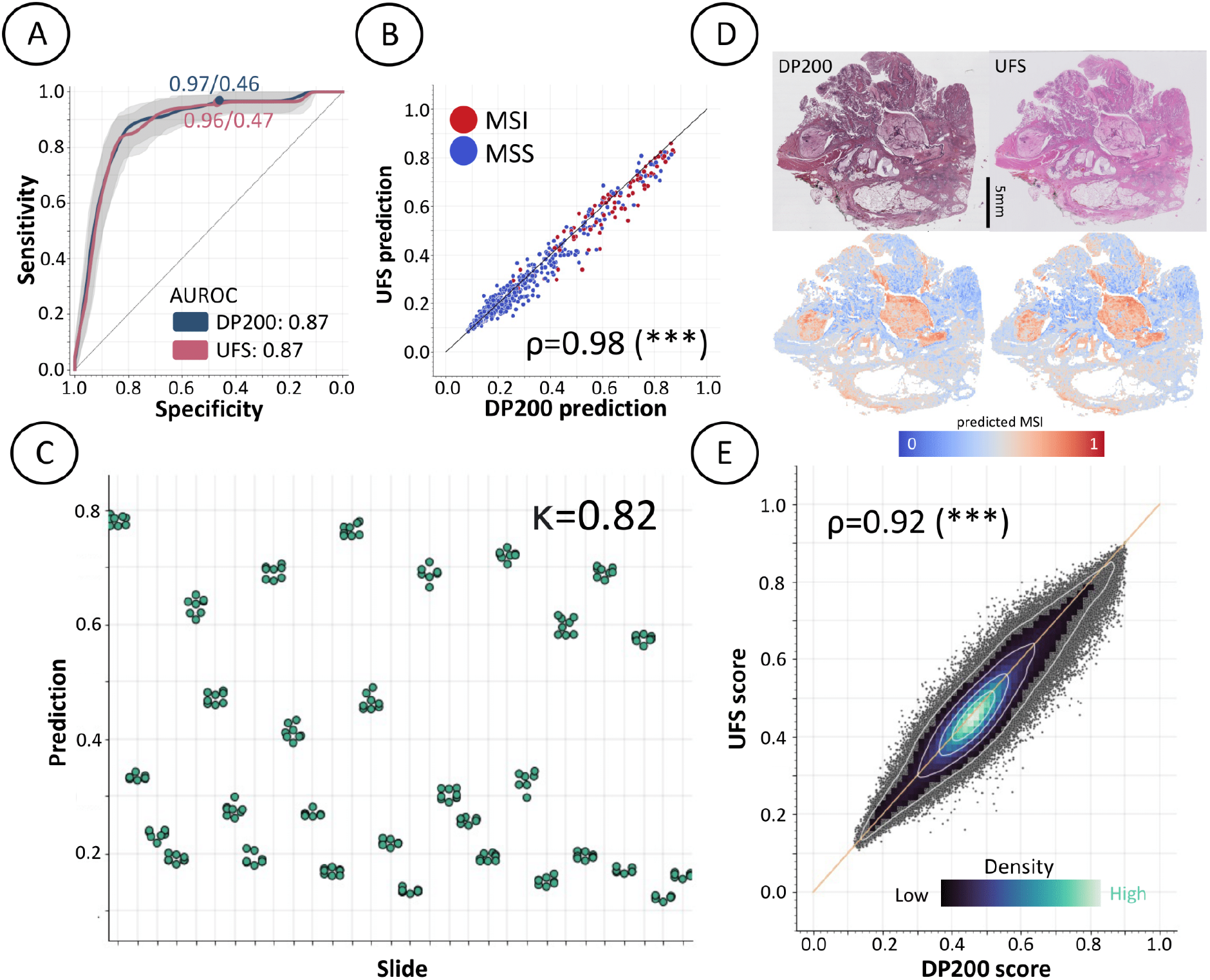
Robustness to scanner variations. **A)** ROC curves of MSIntuit performance on MPATH-DP200 and MPATH-UFS cohorts. To compare performance on the exact same set of patients, we kept the subset of patients that passed QC on the two sets of slides (n=541), and obtained an AUROC of 0.87 on both scanner, **B)** Correlation of the predictions on the same slides on the UFS/DP200 scanners resulting in a Pearson’s correlation of 0.98 (p<0.001), **C)** Prediction distribution for 30 slides, where each slide was digitised 8 times with the UFS scanner. Fleiss’ Kappa of 0.82 was obtained, showing an almost perfect agreement of the tool between the different digitization of the same slide, **D)** Heatmaps showing MSI score for each 112 × 112 μm tile for one representative slide digitised with two scanners, **E)** Correlation of tile MSI scores on DP200 and UFS scanner. MSIntuit outputs a score for each tile, hence we also analysed the concordance of tile scores for a subset of 20 slides digitised with the two scanners (n=272,527 tiles). A Pearson’s correlation of 0.92 was obtained (p<0.001). The colormap representing the spatial density of points indicates that most tile scores were close to the diagonal, showing that tile scores were highly concordant.

### MSIntuit results were consistent across slides obtained from different regions of the tumour

Since several slides are usually available for each patient that may highlight different aspects of the tumour, some criterias are needed to ensure that the slide processed by the tool is representative of the tumour. Guidelines are detailed in the online supplemental material. We showed in the previous section that good performance was obtained with these guidelines. We further explored the consistency of our tool with respect to the region of the tumour processed by digitising additional slides from 1 to 4 other blocks for a subset of 200 out of the 600 tumours of MPATH-DP200 cohort. Average difference of predictions for different slides of the same tumours was low for both MSS and MSI patients with a root mean square error of 0.04 and 0.07, respectively (figure 4B), indicating that the MSIntuit prediction score is consistent between tumour blocks.

**Figure 4.**
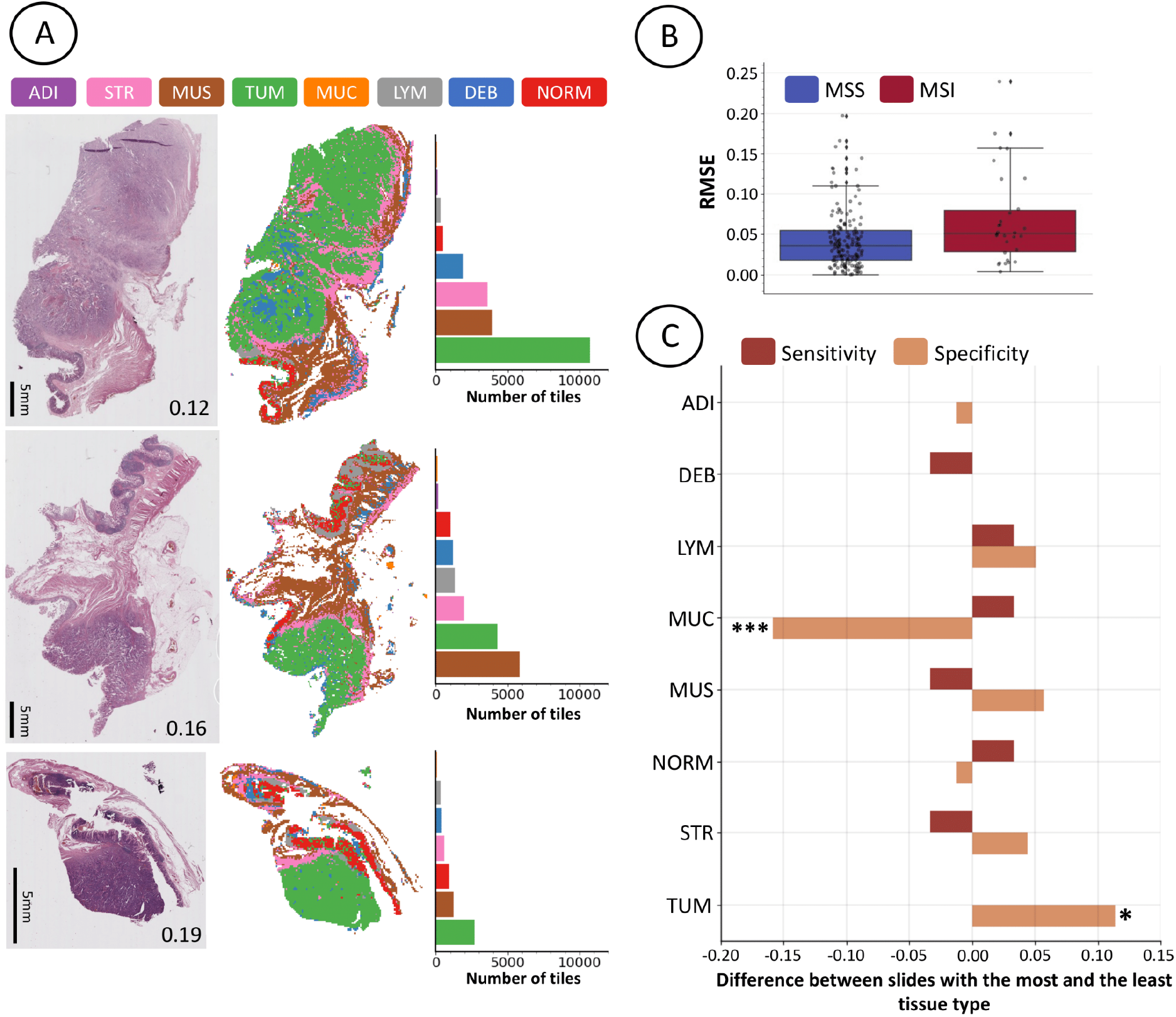
Impact of slide selection on MSIntuit. **A)** Impact of tumour heterogeneity on MSIntuit prediction on a representative pMMR/MSS case. Left: 3 slides picked from different blocks of the same tumour. The number on the bottom right corner of each slide corresponds to the tool’s prediction for the given slide. Middle: segmentation maps using a model trained to categorise tissue into one of the 8 following categories: adipose (ADI), debris (DEB), lymphocytes (LYM), mucus (MUC), smooth muscle (MUS), normal colon mucosa (NORM), cancer-associated stroma (STR), colorectal adenocarcinoma epithelium (TUM). Right: number of tiles belonging to each category. The slide with the largest amount of tumour was the closest to 0; as this patient is MSS, this slide gave the best prediction. **B)** MSIntuit’s predictions variability due to using different slides available for the same patient, for 200 patients with 1 to 4 additional slides of the tumour available. Root mean squared errors (RMSE) of slide prediction and the average of the corresponding patient’ slides were computed and resulted in an average RMSE of 0.04 and 0.07 for MSS and MSI patients respectively, **C)** Difference of sensitivity/specificity when picking slide with the highest and lowest amount of each tissue type was computed for the 200 tumours with multiple slides. Picking the slide with the lowest amount of mucus and largest amount of tumour resulted in a significantly better specificity (+15 points, p<0.001 and +10 points respectively, p<0.05). Other categories were not significantly associated with a better sensitivity or specificity. P-values < 0.05 were considered statistically significant.

For the same set of 200 tumours, we assessed model performance when picking the slide with the highest vs lowest amount of each tissue type, where the amount of tissue of a given type was determined using a ResNet18 model (see section *Impact of tumour heterogeneity analysis* in Materials and Methods). We found that picking the slide with the lowest amount of mucus and largest amount of tumour resulted in a significantly better specificity (+15 points, p<0.001 and +10 points respectively, p<0.05, figure 4C). Other categories were not significantly associated with a better sensitivity or specificity.

### MSIntuit provides interpretable results for pathologists

Four pathologists (T.G., A.A., S.C., J.R.) reviewed the 400 tiles most predictive of MSI (n=200) and non-MSI (n=200) statuses, blinded to their scores. For each tile, the pathologists were asked to annotate the presence of the following histology criteria: normal, fibrosis, inflammation, muscle/vessels, tumour, necrosis, mucus (online supplemental figure 5). Majority voting was used to settle disagreements between pathologists and annotations of a 5th pathologist (D.E.) were used for cases where two pathologists disagreed with the two others. We found that the majority of tiles predictive of both MSI and non-MSI contained tumour cells, with MSI: 70%, non-MSI: 60%). Tiles predictive of MSI were associated with inflammation (MSI: 50%, non-MSI: 13%, p<0.001) and mucus (MSI: 28%, non-MSI: 6%, p<0.001). Tiles predictive of non-MSI were associated with normal glands (MSI: 4%, non-MSI: 26%, p<0.001). These observations are in line with the histological patterns previously described as associated with MSI tumours [20], as well as the interpretability analyses of deep learning models predicting MSI [7,8,21].

## DISCUSSION

With recent CE-IVD approval, MSIntuit is the first AI-based tool that can be used in clinical practice in the EU for MSI pre-screening from an H&E slide of CRC. In this study, we performed a blind validation of this tool, highlighting its value for clinical use. By easily ruling out almost half of the non-MSI population, MSIntuit lightens the workload associated with MMR/MSI testing and enriches the remaining population that need to undergo confirmation screening with MSI patients. This new approach could optimise costs and organisation of MSI testing in pathology labs, especially for countries applying universal MSI screening. The pre-screening approach may also be an opportunity for developing countries where MMR-IHC and MSI-PCR techniques are not accessible or not done systematically due to the costs to screen more patients.

We showed that MSIntuit can serve as a pre-screening tool to rule out almost half of the non-MSI population while correctly classifying more than 95% of dMMR/MSI patients, demonstrating high sensitivity comparable to gold standard methods (92-95%). The blind validation cohort comprised 600 consecutive cases diagnosed in 9 pathology labs in 2017/2018, diminishing the risk of selection bias. Moreover, MSI-PCR was used to confirm doubtful cases for which MMR-IHC analysis was ambiguous to ensure the accuracy of dMMR/MSI labels. It is also important to note that all analyses were pre-specified and that the validation was performed in a one-shot fashion to avoid the risk of overfitting. The model was validated on two different scanners that were not used during training. Altogether, we believe this demonstrates the strength of our validation, as well as the robustness of MSIntuit.

A key technical strength of our approach relies on the use of self-supervised learning to extract features from the histology images. Using this method, we were able to train a feature extractor tailored for histology on 4 million CRC histology images without the need for any labels. As already pointed out by previous studies [22–24], we observed that such feature extractor was more robust to scanner variations and largely outperformed a feature extractor pretrained on ImageNet dataset for MSI prediction task, an approach still widely used in medical imaging.

A key objective of this study was to ensure that the MSIntuit tool could be deployed in clinical practice. To examine the impact of using different scanners at different sites, we digitised 600 slides with two different scanners. We found out that MSIntuit was robust to these variations and reached almost perfect agreement on the two scanners (Cohen’s kappa: 0.82), while its performance was extremely similar with a sensitivity and specificity of 97% / 46% and 96% / 47% on DP200 and UFS scanners respectively. In a recent study, Kleppe highlighted the fact that in the presence of domain shift, sensitivity of a deep learning model could be severely altered. In another study, Echle et al. proposed to use thresholds defined using only training cohorts but the results were mixed, yielding specificities between 15 and 34% [21]. As a mitigation strategy, we developed a calibration approach to ensure that the sensitivity of the MSIntuit tool would be maintained across each new site. We found that this could be accomplished using 30 MSI slides to determine an operating threshold.

Our study has several limitations. MSIntuit was developed and validated solely on slides from surgical specimens. With the recent promising results of NICHE-2 trial [25], neoadjuvant immunotherapy may become the standard of care for CRC patients with dMMR/MSI tumours in the following years. If these results are confirmed, diagnosis of MSI will in the future be predominantly performed on biopsies, while MSIntuit has not been validated on biopsies yet. Echle et al. showed that a model trained to identify MSI on resection specimens transferred well on a cohort of biopsies, which suggests that MSIntuit could also work on biopsies [21]. Further validation on biopsy specimens must be carried out to confirm this hypothesis. Moreover, our tool calibration requires 30 dMMR/MSI slides, which can be difficult to obtain in small centres. Calibration is routinely used for many medical devices, such as MRI, but ideally an AI model should be entirely agnostic to variability in data acquisition across centres.

With the increasing number of biomarkers which should be tested in clinical practice, the need for tools which can ease biomarker testing is greater than ever. With the recent achievements of AI for digital pathology, our tool represents the first step towards the development of AI-based solutions that could identify a panel of actionable biomarkers from a single H&E slide used in clinical routine.

## Data Availability

The discovery TCGA dataset is publicly available at the TCGA data portal (https://portal.gdc.cancer.gov). The external validation PAIP dataset is available at https://paip2020.grand-challenge.org/. Other data produced in the present study are available upon reasonable request to the authors.

## SUPPLEMENTAL

### Supplemental methods

#### Preprocessing of whole-slide images

A preprocessing pipeline was needed to reduce dimensionality and clean the data before training any model. The first step of our pipeline consisted of detecting the tissue on the WSI: a U-Net neural network [26] was used to segment part of the image that contains relevant matter, and discard artefacts such as blur, pen marker etc., as well as the background. This U-Net network was previously trained on 460 H&E and MMR-IHC slides from an internal dataset where tissue was manually annotated, and validated on 115 slides with a Dice score of 0.96. This network was applied on images of size 2048 × 2048 μm (512 × 512 px, at a resolution of 4 MPP) extracted from the WSI. The second step consisted of splitting the slide into smaller images, called “tiles”, of 112 × 112 μm (224 × 224 px, at a resolution of 0.5 MPP). At least 50% of the tile must have been detected as foreground by the U-Net model to be kept. For training, a maximum of 8,000 tiles were extracted from each slide while all tiles were extracted for inference. The final step consisted of extracting features from each tile: 2,048 relevant features were extracted using a wide Resnet50 network [27] (the bottleneck number of channels is twice larger in every block) trained in a self-supervised fashion with MoCo v2 [28]. This network was trained on 4 million tiles from the TCGA-COAD dataset, with massive data augmentation (random cropping, random flips, colour jitter, random grayscale, gaussian blur), and without using any labels. Feature extractor weights were frozen both for inference and training.

#### Slide selection

Guidelines regarding slide selection defined to guide pathologists for the use of MSIntuit in clinical practice were to follow the maximum number of the following criteria: the slide with the largest surface of tumour tissue, the slide with the most invasive tumour, the slide with the least necrosis, the slide must not contain preparation artefacts (staining artefacts, folds on the fabric cut, residual air or water bubbles, traces of marker, damaged coverslips, scanning artefacts).

#### Bland-Altman plot to assess inter-scanner reliability

The Bland-Altman plot was also used (online supplemental figure 4) to assess the agreement between DP200 and UFS prediction scores, and the 95% limits of agreement (LoA) were calculated as mean±1.96 standard deviation (SD) of the difference (DP200 Score - UFS score). A p-value < 0.05 was considered statistically significant.

#### Interpretation of ICC and Cohen’s Kappa values

An ICC below 0.5 indicates poor reliability, an ICC between 0.5 and 0.75 indicates moderate reliability, an ICC between 0.75 and 0.9 indicates good reliability, and an ICC above 0.9 indicates excellent reliability [29]. A Cohen’s kappa under 0.2 indicates slight agreement, 0.21 to 0.40 indicates fair agreement, 0.41–0.60 indicates moderate agreement, 0.61–0.80 indicates substantial agreement, and 0.81 to 1.0 indicates almost perfect agreement [30].

#### Slide registration

WSIs of the samples obtained with the DP200 and UFS scanners were not perfectly aligned because of each scanner’s principles of operations (orientation of the objective, automatic cropping of empty regions, etc). To compare tile individual scores across the two scanners (figure 2E), we therefore used an image registration process to make sure the local regions of one slide match the local regions of its counterpart digitised with the other scanner. This registration process was done using the Elastix and Transformix softwares [31,32]. Non-rigid registration parameters were first computed on sub-sampled WSIs (8μm per pixel), optimising the Mattes Advanced Mutual Information on ten consecutive levels of resolution. Those parameters were finally applied to the high resolution UFS WSI in order to obtain aligned WSIs at identical resolutions.

## Supplementary figures

**Supplemental figure 1.**
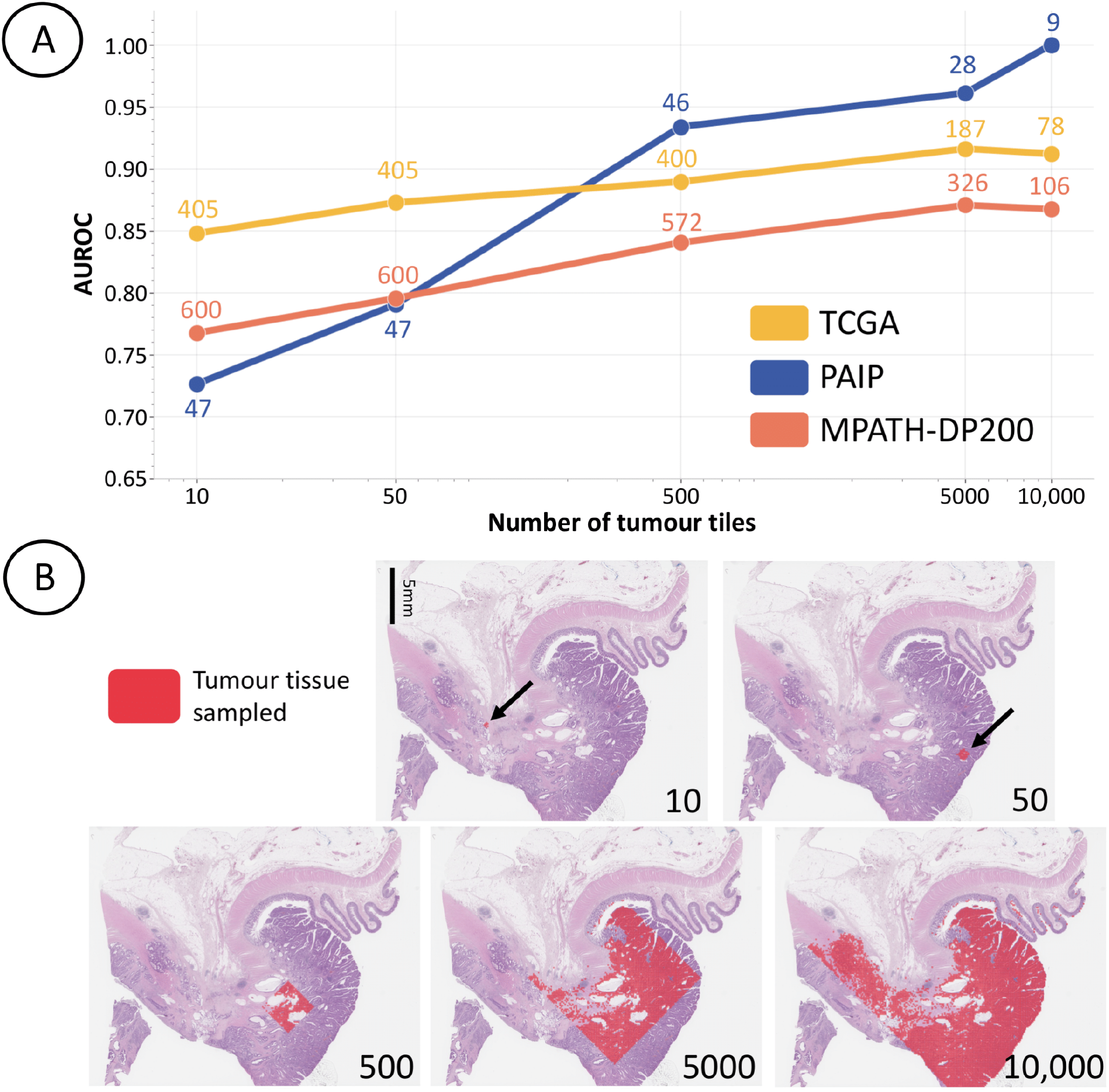
Impact of amount of tumour on the model. To assess the minimum amount of tumour on the slide needed to ensure MSIntuit yields good performance, we looked at how the number of tumour tiles impact the results obtained on TCGA and PAIP cohorts before performing the blind-validation. **A)** For a number *x* being 10, 50, 500, 5000, 10000, we randomly selected an area of x tumour tiles for each slide and performed the prediction on it. Slides with less than x tumour tiles were discarded. Number of slides that contain at least x tumour tiles are displayed next to each point. X-axis is in log scale, **B)** Example of tumour areas selected, for different numbers of tumour tiles (bottom right corner of each image).

**Supplemental figure 2.**
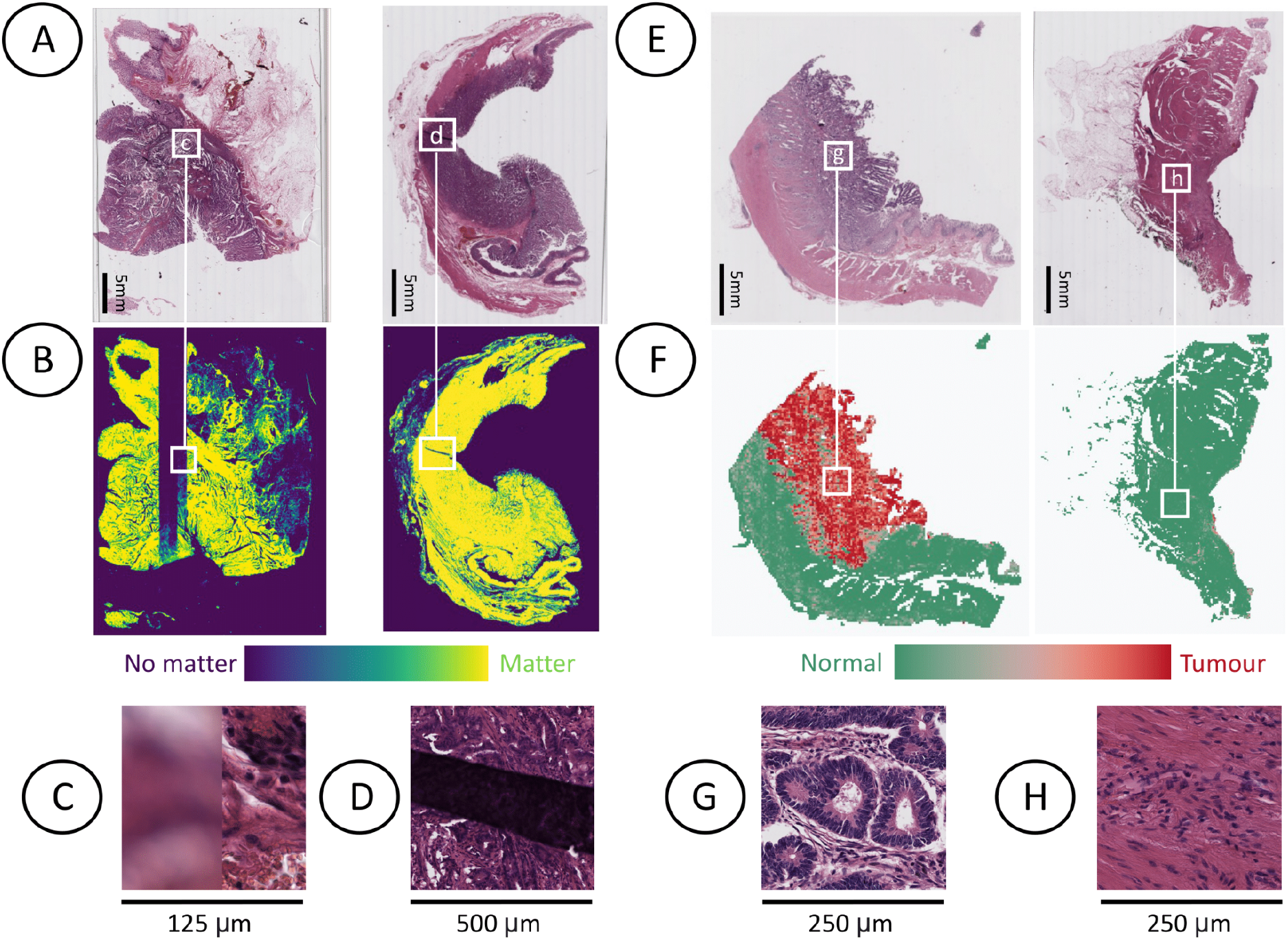
Quality Check. **A)** Left: slide with a blurry strip due to a digitization issue, not noticeable at low resolution, right: slide with a tissue fold. **B)** Matter detection heatmaps of the UNet neural network integrated in MSIntuit’s preprocessing and QC procedures. Blurry regions (left) and tissue fold (right) are not detected as matter. **C), D)** Zoomed-in images of blurry and tissue fold regions. **E)** Slide with abundant tumour tissue that passed QC (left), slide with too few tumour tissue (<500 tumour tiles) that did not pass QC. **F)** Corresponding tumour heatmaps obtained with a tumour classifier part of MSIntuit’s QC procedure. **G), H)**, Zoomed-in images of tumour (left) and (normal) regions of left and right slide, respectively.

**Supplemental figure 3.**
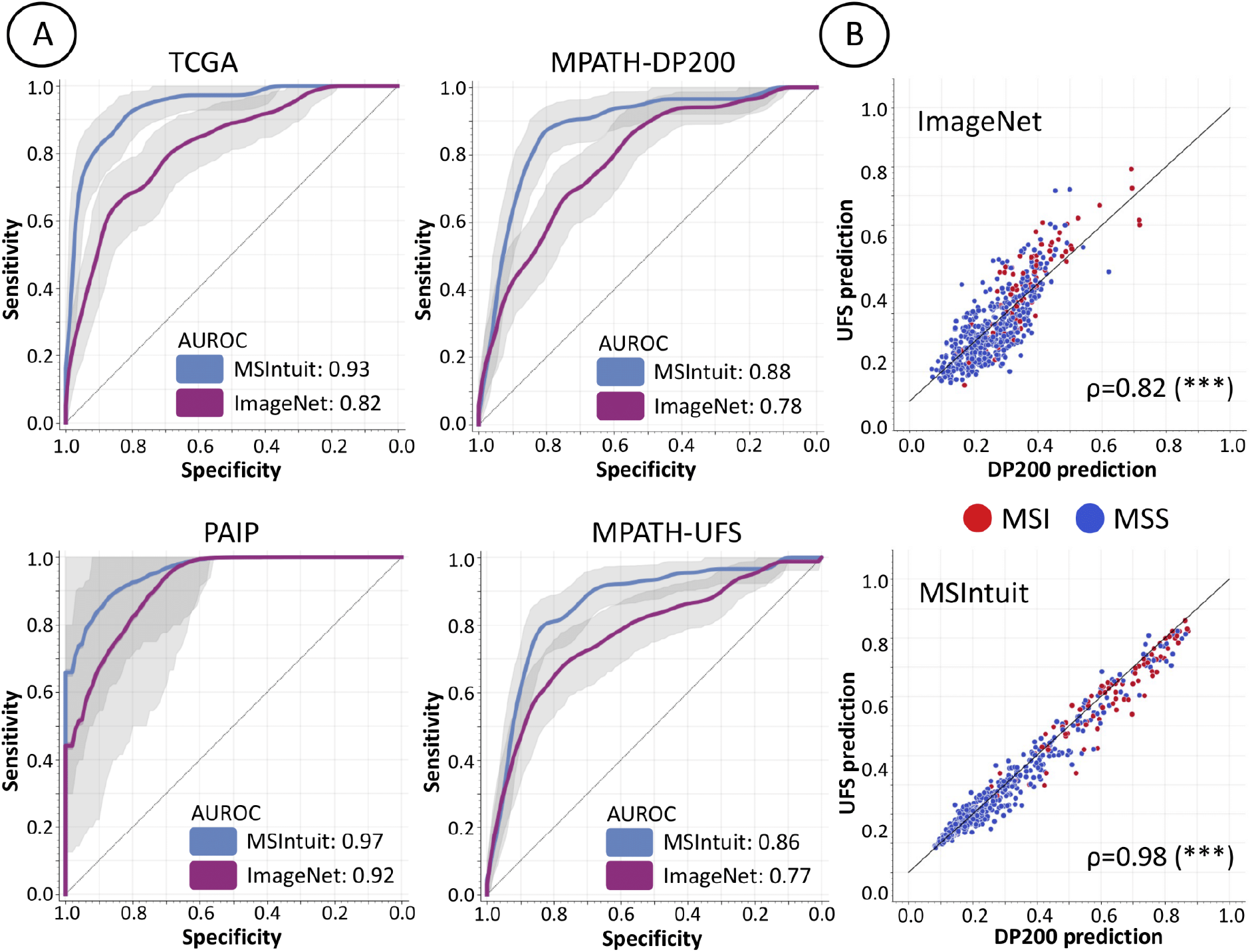
Comparison of Self-Supervised Learning and ImageNet-pretrained approaches. **A)** ROC curves of our tool’s method which uses a feature extractor trained with MoCo v2 on millions of colorectal cancer images (MSIntuit) and a method which uses a feature extractor trained on ImageNet dataset in a supervised fashion (ImageNet), on TCGA, PAIP, MPATH-DP200 and MPATH-UFS cohorts. Apart from the feature extraction part, the same pipeline was used for the two methods (QC, calibration, downstream model etc..). **B)** Correlation of the predictions on the same slides on the UFS/DP200 scanners for ImageNet and MSIntuit methods resulted in a Pearson’s correlation of 0.82 (p<0.001) and 0.98 (p<0.001) for ImageNet and MSIntuit methods, respectively.

**Supplemental figure 4.**
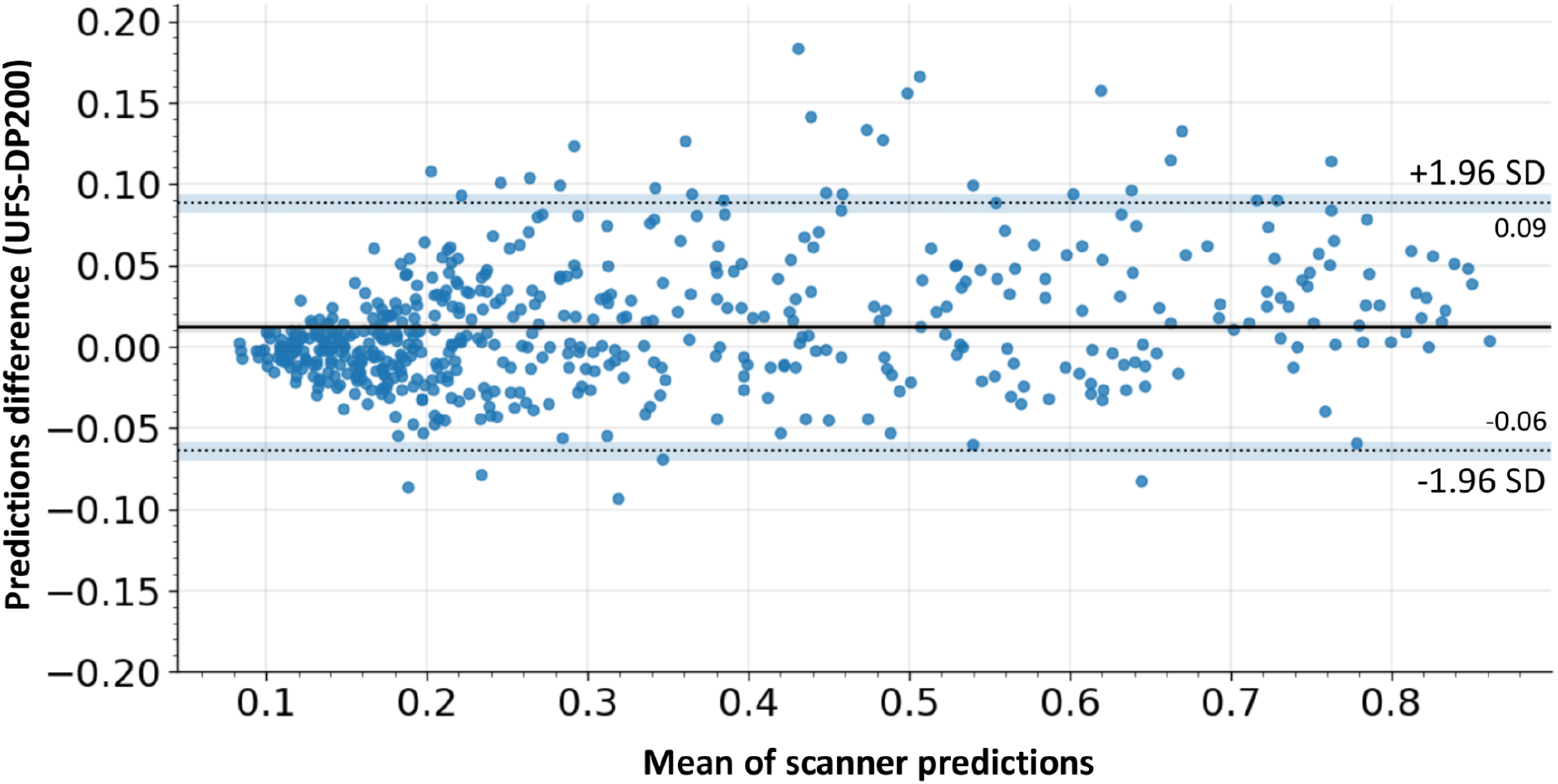
Bland-Altman plot for inter-scanner reliability. A Bland-Altman plot to analyse the agreement of MSIntuit predictions on UFS and DP200 scanners by looking at the mean inter-scanner difference of prediction scores. A relatively low prediction score variability was observed with an overall mean inter-scanner score difference of 0.01 (where the MSIntuit score can vary between 0 and 1) with a limit of agreement 95% confidence interval ranging from −0.06 to 0.09.

**Supplemental figure 5.**
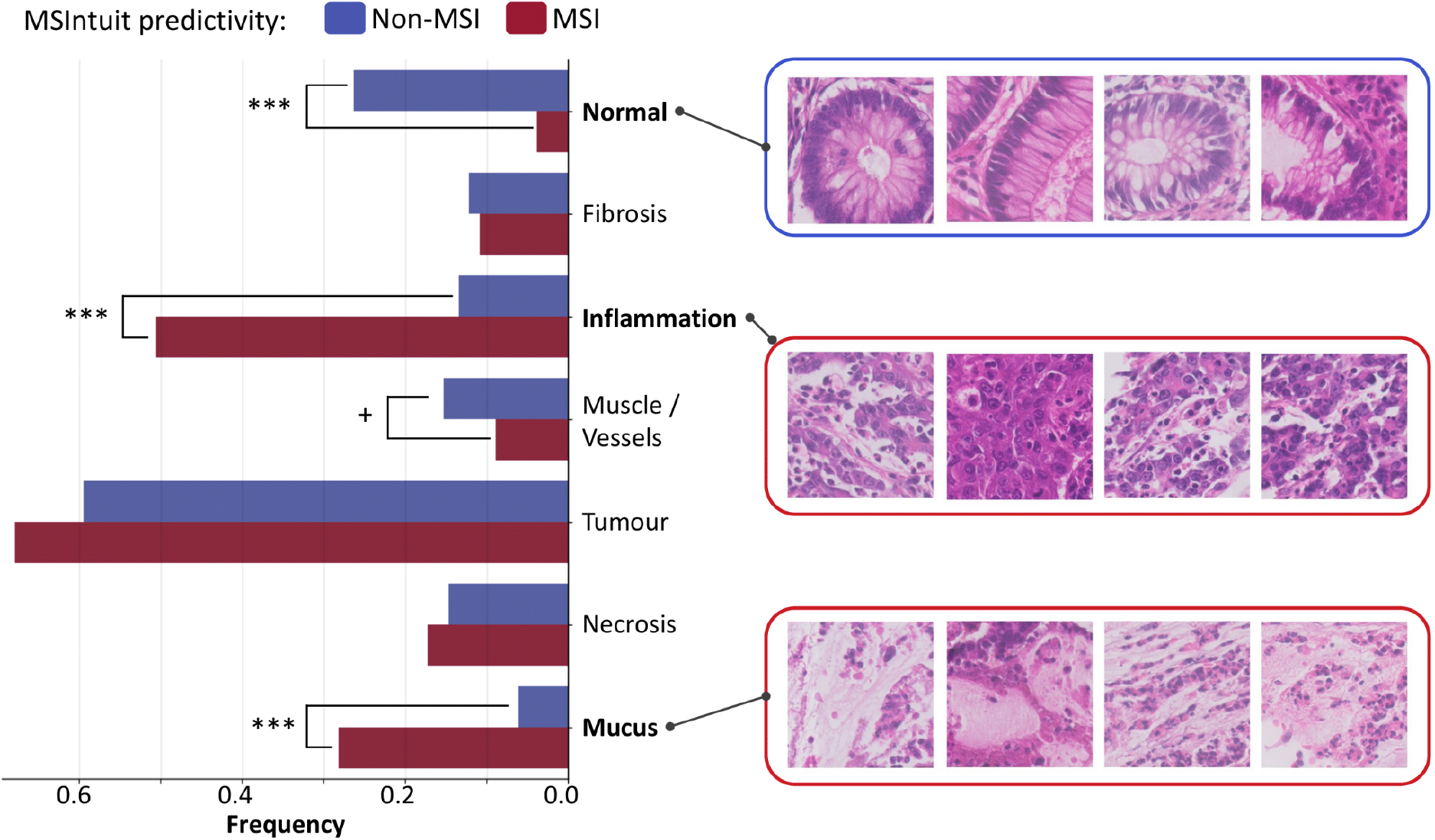
Interpretability analysis. Proportion of histology patterns associated with non-MSI and MSI according to MSIntuit. Four pathologists reviewed the 400 tiles most predictive of MSI (n=200) and non-MSI (n=200) statuses, blinded to their scores. Majority of tiles predictive of both MSI and non-MSI contained tumour cells, with MSI: 70%, non-MSI: 60%). Tiles predictive of MSI were associated with inflammation (MSI: 50%, non-MSI: 13%, p<0.001) and mucus (MSI: 28%, non-MSI: 6%, p<0.001). Tiles predictive of non-MSI were associated with normal glands (MSI: 4%, non-MSI: 26%, p<0.001).

**Supplemental figure 6.**
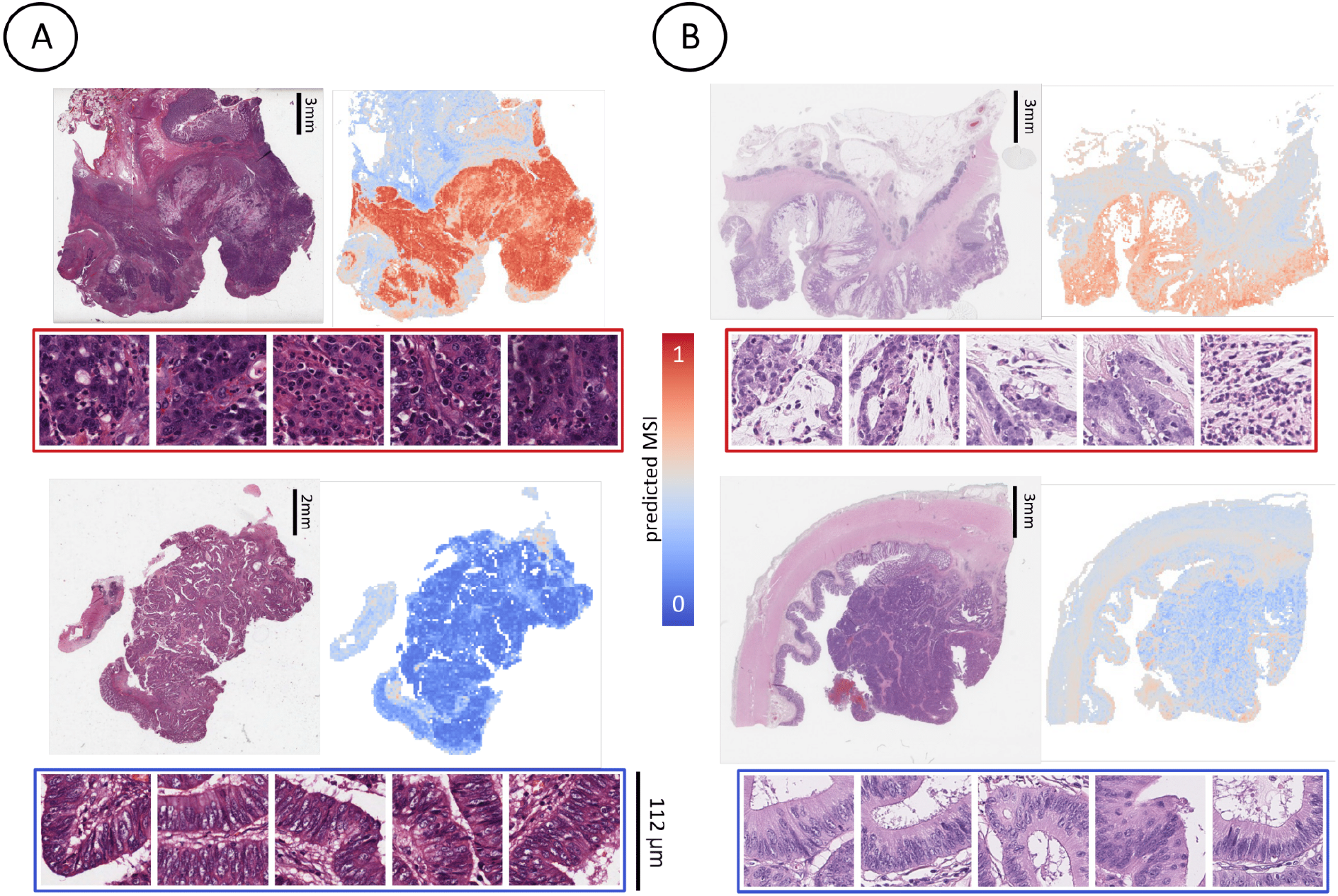
Model’s interpretability on TCGA & PAIP cohorts.Heatmaps of the tool with corresponding most predictive tiles of a representative dMMR/MSI case (top) and a pMMR/MSS case (bottom) of. **A)** TCGA cohort, **B)** PAIP cohort.

### Supplemental tables

**Supplemental table 1.** Cohorts description.

|  | TCGA | PAIP | Medipath<br>(MPATH-DP200/MPATH-UFS) |
| --- | --- | --- | --- |
| Number of patients | 434 | 47 | 600 |
| Region | United States | South Korea | France |
| H&E FFPE slides, n | 427 | 47 | 600 |
| H&E Frozen slides, n | 432 | - | - |
| MSI patients, n (%) | 78 (18%) | 12 (26%) | 123 (21%) |
| dMMR/MSI diagnosis | MSI-PCR | MSI-PCR | MMR-IHC 4-plex, followed by MSI-PCR for indeterminate cases |
| Scanner | Aperio | Aperio AT2 | Ventana DP200 & Phillips Ultra Fast Intellisite |
| Age at diagnosis, IQR | 68 (58-77) | - | 74 (64-82) |
| Grade 1, n (%) | - | - | 219 (39%) |
| Grade 2, n (%) | - | - | 296 (53%) |
| Grade 3, n (%) | - | - | 46 (8%) |
| Stage I, n (%) | 70 (17%) | - | 46 (8%) |
| Stage II, n (%) | 157 (39%) | - | 91 (15%) |
| Stage III, n (%) | 117 (29%) | - | 281 (47%) |
| Stage IV, n (%) | 60 (15%) | - | 162 (27%) |

**Supplemental table 2.** Performance comparison using SSL pre-trained and ImageNet pre-trained feature extractors. We compared the results of two methods: the first one being the one of MSIntuit which uses a feature extractor trained with MoCo v2 on millions of colorectal cancer images, while the second one uses a feature extractor trained on ImageNet dataset in a supervised fashion. Apart from the feature extraction, the same pipeline was used for the two methods (QC, calibration, downstream model etc..). Results obtained on TCGA (cross-validation), PAIP and MPATH-DP200 cohorts are reported.

| Cohort | Metric | ImageNet 3rd block | ImageNet Last block | MoCo v2 (MSIntuit) |
| --- | --- | --- | --- | --- |
| TCGA | AUROC | 0.80 +- 0.05 | 0.81 +- 0.04 | **0.93 +- 0.03** |
| PAIP | AUROC | 0.92 [0.84-0.97] | 0.88 [0.73-0.98] | **0.97 [0.90-0.99]** |
| MPATH-DP200 | AUROC | 0.78 [0.74-0.83] | 0.77 [0.72-0.82] | **0.88 [0.84-0.91]** |
|  | Sensitivity | 0.94 [0.90-0.98] | 0.92 [0.87-0.96] | **0.97 [0.93-0.99]** |
|  | Specificity | 0.31 [0.27-0.34] | 0.31 [0.28-0.35] | **0.46 [0.42-0.5]** |
| MPATH-UFS | AUROC | 0.77 [0.72-0.82] | 0.72 [0.67-0.77] | **0.86 [0.83-0.90]** |
|  | Sensitivity | 0.92 [0.87-0.97] | 0.85 [0.78-0.91] | **0.95 [0.90-0.98]** |
|  | Specificity | 0.28 [0.24-0.31] | 0.36 [0.33-0.4] | **0.47 [0.43-0.51]** |

**Supplemental table 3.** Training the model on FFPE slides only versus FFPE and frozen slides of TCGA-COAD. Both FFPE and snap-frozen slides are available for most patients of the TCGA-COAD dataset, the dataset we used for training. Although MSIntuit is intended to be used on FFPE slides, we found that using frozen slides in addition to FFPE ones during Chowder training slightly improved performance when validating the tool on FFPE samples, likely because the Chowder model gained robustness with this augmentation strategy all the while doubling our sample size. In the table below, we compared the performance of two models: one model trained using only FFPE slides, and another model which uses both FFPE and frozen slides for training (MSIntuit). In the table below, we display the results obtained when validating on FFPE slides of TCGA-COAD (cross-validation), PAIP and MPATH-DP200 datasets (external validation).

| Cohort | Metric | FFPE Only | FFPE & Frozen (MSIntuit) |
| --- | --- | --- | --- |
| TCGA-COAD | AUROC | 0.91 +- 0.02 | **0.93 +- 0.03** |
| PAIP | AUROC | 0.97 [0.91-1.00] | 0.97 [0.91-1.00] |
| MPATH-DP200 | AUROC | 0.87 [0.83-0.90] | **0.88 [0.84-0.91]** |
|  | Sensitivity | 0.93 [0.88-0.97] | **0.97 [0.93-0.99]** |
|  | Specificity | **0.57 [0.53-0.60]** | 0.46 [0.42-0.5] |
| MPATH-UFS | AUROC | 0.86 [0.82-0.89] | 0.86 [0.83-0.90] |
|  | Sensitivity | 0.95 [0.90-0.98] | 0.95 [0.90-0.98] |
|  | Specificity | 0.42 [0.38-0.46] | **0.47 [0.43-0.51]** |

**Supplemental table 4.** Training/Testing on tumour regions only. Even though known MSI-related features are found only within tumour regions, we found that applying our model on the whole slide yielded slightly better results. In the table below, we compare the performance of two models: one model trained and validated using only tumour regions of the slide, and MSIntuit which keeps the whole slide for training and validation. tumour regions were defined using a tumour detection model (see section Quality Checks of Material and Methods).

| Cohort | Metric | Tumour Only | Whole slide (MSIntuit) |
| --- | --- | --- | --- |
| TCGA-COAD | AUROC | 0.90 +- 0.03 | **0.93 +- 0.03** |
| PAIP | AUROC | 0.94 [0.85-0.99] | **0.97 [0.90-0.99]** |
| MPATH-DP200 | AUROC | 0.87 [0.84-0.90] | **0.88 [0.84-0.91]** |
|  | Sensitivity | 0.95 [0.90-0.98] | **0.97 [0.93-0.99]** |
|  | Specificity | 0.44 [0.40-0.47] | **0.46 [0.42-0.5]** |

**Supplemental table 5.** Performance of MSIntuit repeating threshold decision procedure. Since the calibration step involves selecting some slides to define an appropriate operating threshold, we analysed how the selection of these slides may impact the model performance. To this end, we repeated the calibration step 1000 times (selecting each time a different set of slides to calibrate the tool, and assessing the performance of the model on the remaining patients).Metrics obtained with this experiment are reported in the table below.

|  | MPATH-DP200 | MPATH-UFS |
| --- | --- | --- |
| AUROC | 0.88 [0.87-0.89] | 0.86 [0.85-0.88] |
| Sensitivity | 0.94 [0.83-1.0] | 0.94 [0.84-1.0] |
| Specificity | 0.52 [0.16-0.80] | 0.47 [0.14-0.72] |

**Supplemental table 6.**
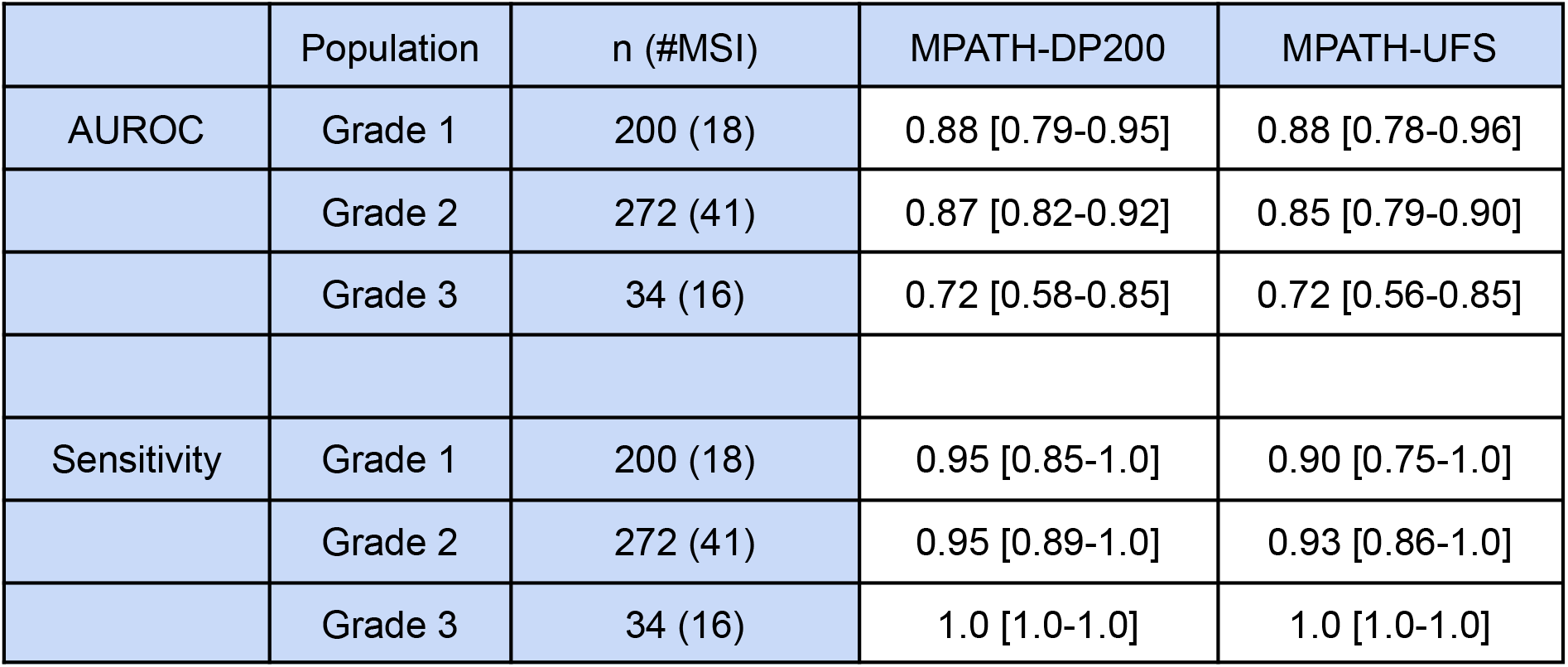
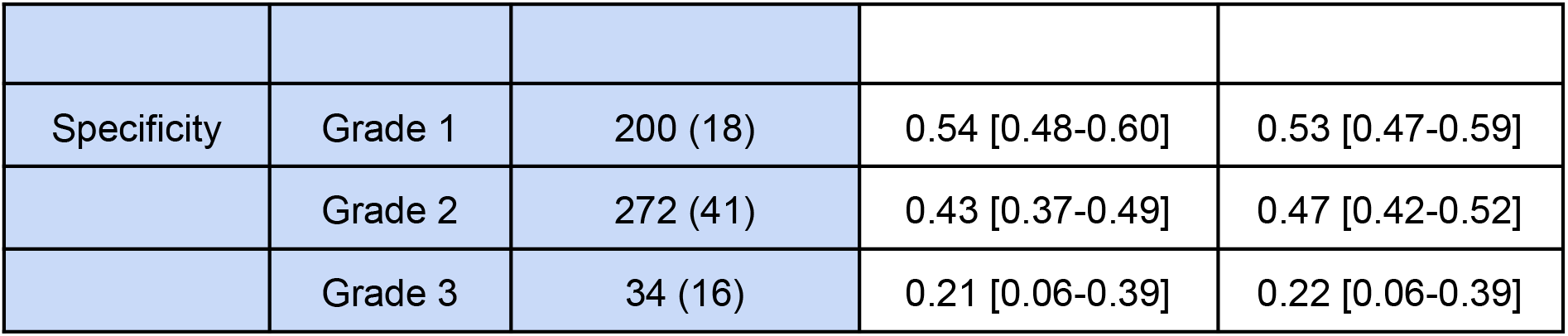
Performance of MSIntuit inside histological grade subgroups. We looked at the tool’s performance on subgroups of MPATH-DP200 and MPATH-UFS cohorts with histological grade 1, 2 or 3.

|  | Population | n (#MSI) | MPATH-DP200 | MPATH-UFS |
| --- | --- | --- | --- | --- |
| AUROC | Grade 1 | 200 (18) | 0.88 [0.79-0.95] | 0.88 [0.78-0.96] |
|  | Grade 2 | 272 (41) | 0.87 [0.82-0.92] | 0.85 [0.79-0.90] |
|  | Grade 3 | 34 (16) | 0.72 [0.58-0.85] | 0.72 [0.56-0.85] |
| Sensitivity | Grade 1 | 200 (18) | 0.95 [0.85-1.0] | 0.90 [0.75-1.0] |
|  | Grade 2 | 272 (41) | 0.95 [0.89-1.0] | 0.93 [0.86-1.0] |
|  | Grade 3 | 34 (16) | 1.0 [1.0-1.0] | 1.0 [1.0-1.0] |
| Specificity | Grade 1 | 200 (18) | 0.54 [0.48-0.60] | 0.53 [0.47-0.59] |
|  | Grade 2 | 272 (41) | 0.43 [0.37-0.49] | 0.47 [0.42-0.52] |
|  | Grade 3 | 34 (16) | 0.21 [0.06-0.39] | 0.22 [0.06-0.39] |

**Supplemental table 7.**
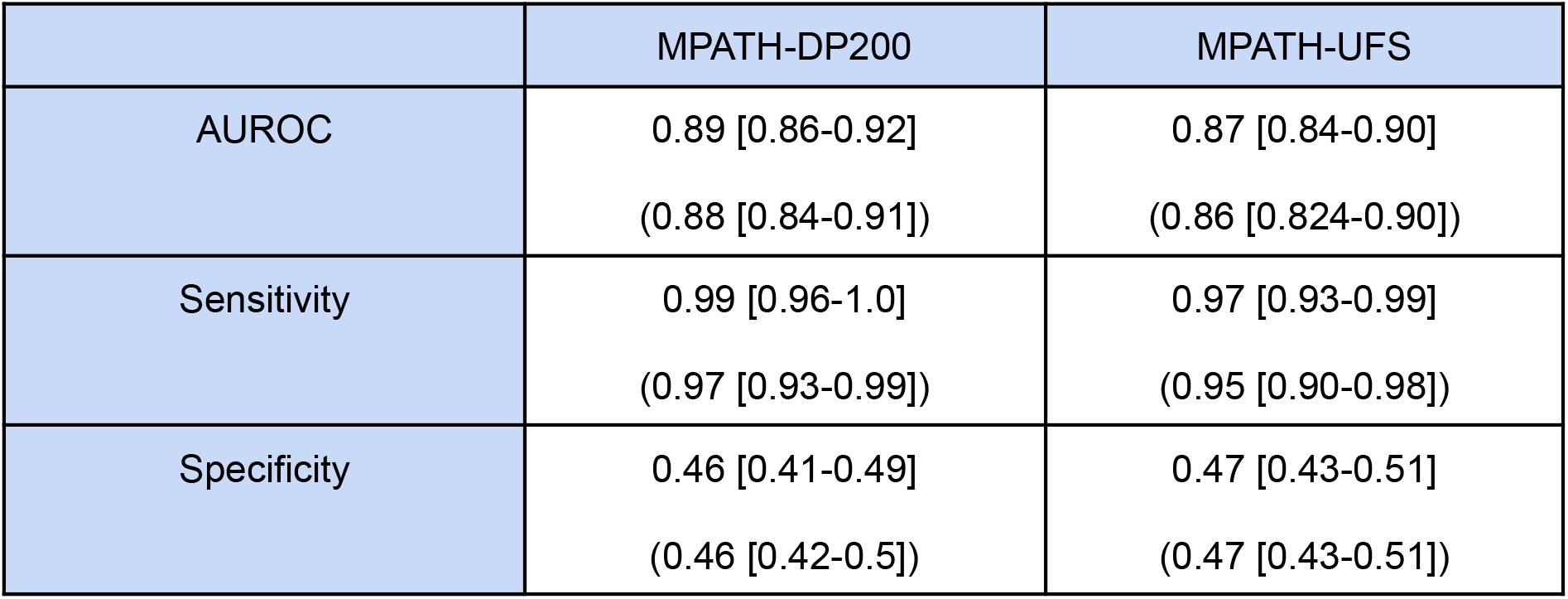
Performance of MSIntuit removing patients with *rare* protein loss. We define rare protein loss as one of the following protein losses: loss of MLH1 without loss of PMS2, loss of MSH2 without loss of MSH6, loss of MLH1 and loss of MSH2[33]. In the table below, we report the performance of MSIntuit in the population of patients that did not have a rare protein loss. Numbers in brackets represent 95% confidence intervals, generated using bootstrap with 1,000 repeats. Numbers in parentheses represent metrics obtained on the whole population.

**Supplemental table 8.**
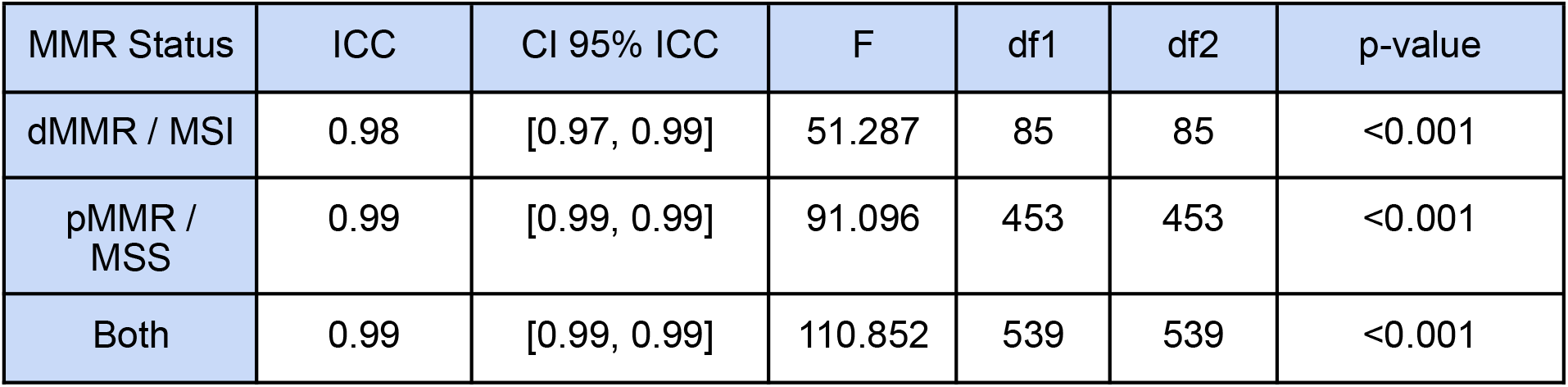
Intraclass Correlation Coefficient (ICC). *F*: value of the F-test, *df*: degrees of freedom. We analysed inter-scanner reliability by computing the ICC scores. An F-test is performed in order to confirm or not the presence of bias during ICC computation. It is computed as the ratio of the mean square error between measurements over the total mean squared error. The degrees of freedom are an indication of the total number of subjects used in the analysis. As suggested by Liljequist et al.[14], an F-value considerably smaller than the total sample size indicates that biases are weak.

## Acknowledgments

The results published here are in whole or part based upon data generated by the TCGA Research Network: https://www.cancer.gov/tcga. Regarding the PAIP dataset: De-identified pathology images and annotations used in this research were prepared and provided by the Seoul National University Hospital by a grant of the Korea Health Technology R&D Project through the Korea Health Industry Development Institute (KHIDI), funded by the Ministry of Health & Welfare, Republic of Korea (grant number: HI18C0316).

This work was granted access to the HPC resources of IDRIS under the allocation AD011012519 made by GENCI.

We thank Pierre Courtiol, Simon Jégou, Benoit Schmauch and Olivier Moindrot for their contribution in the early development of the model. We thank Sanjana Vasudevan for her corrections of the manuscript and Sarah Ulstrup for her help in designing the figures. We also thank Dr Alicia Tourneret, Dr Damienne Declerck, Céline Coppolani, Pauline Mespoulhe and Caroline Rancati for their help in collecting data of the validation cohort.

## Contributors

Study conception and design: CS, AF, M. Sefta, MA; data collection: TG, AA, SC, JR, DE, SR; Software: CS, RD, OT, NL; analysis and interpretation of results: CS, RD, OT, TG, AA, SC, JR, DE, M. Svrcek, JNK; draft manuscript preparation: CS, AF, LG, OT. All authors reviewed the results and approved the final version of the manuscript.

## Patient consent for publication

Not applicable.

## Ethics approval

All experiments were conducted in accordance with the General Data Protection Regulation (GDPR) and the French laws and regulations. The Medipath data subjects have generally been informed for the re-use of their samples and data collected during the care for research purposes. Medipath has obtained an approval of the “Ministère de l’Enseignement Supérieur, de la Recherche et de l’Innovation (MESRI)” for the storage of samples for research purposes and has nominatively reinformed patients for the reuse of their data for Owkin’s experiments.

